# Living in Endemic Area for Infectious Diseases is Associated to Differences in Immunosenescence Signatures

**DOI:** 10.1101/2024.12.16.24319110

**Authors:** Monique Macedo Coelho, Felipe Caixeta Moreira, Luciana Werneck Zuccherato, Lucas Haniel de Araújo Ventura, Giovanna Caliman Camatta, Bernardo Starling-Soares, Lícia Torres, Danielle Fernandes Durso, Hugo Itaru Sato, Murilo Soares da Costa, Henrique Cerqueira Guimarães, Rafael Calvão Barbuto, Mauro Lúcio O. Júnior, Elaine Speziali, Unaí Tupinambas, Santuza Maria Ribeiro Teixeira, Gabriela Silveira-Nunes, Andrea Teixeira-Carvalho, Tatiani Uceli Maioli, Ana Maria Caetano Faria

## Abstract

Research on aged individuals from developed countries show that lifestyle factors such as diet, physical activity, stress, smoking, and sleep quality impact aging. However, other relevant factors may influence aging in less-studied populations, such as Brazilian cohorts. This study aimed to analyze immunosenescence profile of individuals living in an endemic area for several infectious diseases in Brazil. We showed that these individuals exhibited accelerated epigenetic aging and increased production of IL-12p70, IL-17A, and IL-9. Production of inflammatory mediators IL-12p70, IL-6, IL-1β, IL-2, and IL-1ra in individuals with flu-like symptoms and those with COVID-19 was higher among residents in endemic areas than in residents from a control non-endemic area. Furthermore, residents of the endemic area had a more prominent inflammatory profile during viral infection and a different pattern of plasma mediators when compared to residents of a non-endemic area. It suggests that these two cohorts had specific immune signatures regardless of the presence or the type of infection at study. Therefore, we demonstrated that there were distinct patterns of immune responses and epigenetic aging depending on the environment the individuals live in. These observations add a layer of diversity to the studies of human aging by including less represented individuals.

## Introduction

Aging is a multifactorial and heterogeneous biological phenomenon that affects individuals in diverse ways(Cohen et al., 2020). Lifestyle factors, such as nutritional habits, physical activity, well-being, stress, smoking, and sleep quality, significantly influence the aging process(Bloomberg et al., 2024; Friedman, 2020).

Aging affects all body compartments, and the immune system is one of the most affected. Changes in immune activity are known as immunosenescence and it includes a chronic state of low-grade inflammation, termed “inflammaging,” characterized by alterations in the levels of plasma mediators such as IL-6, TNF-α, IL-1, and IL-10(FRANCESCHI et al., 2000; Fulop et al., 2018). Inflammaging is a systemic consequence of various aging-related events, including epigenetic changes, mitochondrial dysfunction, genomic instability, and the emergence of the senescence-associated secretory phenotype (SASP) by senescent cells(Franceschi et al., 2007).

Immunosenescence increases the susceptibility to a range of diseases(Hanlon et al., 2018; Kennedy et al., 2014). However, the modifications that accompany aging are often compensatory and protective for living organisms. Grosse and colleagues demonstrated that senescent cells play important physiological roles, and their elimination could lead to health deterioration and a shorter lifespan in mice due to the disruption of blood-tissue barriers(Grosse et al., 2020). Moreover, not all aged individuals suffer from chronic inflammatory diseases and frailty. Studies on healthy nonagenarians and centenarians in Italy and in healthy elderly people in Brazil showed that these individuals develop remodelling mechanisms that control the deleterious effects of inflammation and immunosenescence(Caetano Faria et al., 2008; Caruso et al., 2022; Lima-Silva et al., 2024; Santoro et al., 2021).

In a previous study, we reported that the response triggered by continuous antigenic stimuli can lead to accelerated senescence in individuals residing in endemic areas(Durso et al., 2022). Herein, we examined aging in an endemic area (EA) in Brazil, where the prevalence of well-characterized infections such as schistosomiasis, leishmaniasis, and leprosy is high. Immunosenescence has been known as a consequence of chronic antigenic exposure(De Martinis et al., 2005). However, there is no detailed evaluation of the immunological repercussions in the immunosenescence profile of individuals residing in EAs compared to those living in non-endemic areas (NEAs).

DNA methylation is one of several mechanisms that regulate gene expression and is considered an important biomarker of aging. Based on methylation status, epigenetic clocks have been developed to calculate individuals’ biological age. These clocks can detect not only the acceleration of aging but also the effects of deceleration in models of healthy aging(Field et al., 2018).

Moreover, DNA methylation patterns are associated with a wide range of age-related diseases, including Alzheimer’s(Pellegrini et al., 2021), cardiovascular diseases(Fernández-Sanlés et al., 2021), and cancer (Durso et al., 2017). Age acceleration measured by methylation patterns are known to be altered in response to environmental stimuli such as exercise (Turner et al., 2020), diet (Pauwels et al., 2017), smoking (Joubert et al., 2016), and pollutants (Plusquin et al., 2017). One study demonstrated that an eight-week treatment program, including diet, sleep, exercise, relaxation techniques, and probiotic supplementation, was able to reduce the epigenetic age of a group of 43 adult men by 3.23 years (Fitzgerald et al., 2021).

Traditionally aging studies focuses on cohorts and issues associated with developed countries and Caucasian populations. Our hypothesis in this study is that, in addition to lifestyle, other significant environmental factors such as chronic exposure to infectious agents may be associated with distinct patterns of immunosenescence and aging. Therefore, in this study, we aim to evaluate the immunological repercussions in individuals residing in a cohort of individuals from an endemic area in Brazil.

## Material and methods

### Ethics statement

This study meticulously adhered to the ethical standards set forth by National Research Ethics Committee (CONEP), prioritizing the safety and well-being of all participants. It was approved by Human Research Ethical Board of UFMG (CEP-UFMG) and the National Research Ethics Committee (CONEP) (CAAE # 40208320.3.1001.5149). All volunteers were informed of the objectives and procedures involved in the study and they signed the Informed Consent Form (TCLE) before their voluntary participation.

### Study subjects

The research was conducted in two distinct Brazilian cities: one living in a non-endemic area for infectious diseases (Belo Horizonte/MG), where 107 participants were enlisted, and another in an endemic region (Governador Valadares/MG), also with 107 participants. This amassed a cohort of 214 individuals, whose attributes are elaborated upon in Table 1.

**Table 1.** Characterization of the study sample:

|  | AE (endemic area) |  |  | NEA (non-endemic area) |  |  |
| --- | --- | --- | --- | --- | --- | --- |
|  | Flu- Like<br>n = 45 | COVID-19<br>n = 47 | Negative<br>Control<br>n = 15 | Flu- Like<br>n = 45 | COVID-19<br>n = 47 | Negative<br>Control<br>n = 15 |
| Age<br>(years) | 38 (18-60) | 38,5 (18-77) | 41 (19-78) | 36 (19-63) | 40,5 (18-76) | 39 (18-65) |
| Sex Female<br>(%) | 51,1 | 46,8 | 46,7 | 46,6 | 53,19 | 53,3 |
| Sex Male (%) | 48,9 | 54,2 | 53,3 | 53,4 | 46,80 | 46,6 |
| BMI (Kg/m<sup>2</sup>) | 28,3 ±5,57 | 26,29 ±4,53 | 26 ±4,21 | 28,4 ±7 | 30 ±5,67 | 27 ±6,82 |
| Viral Load<br>(ct) | – | 21,89 ±7,12 | – | – | 27,3 ±5,14 | – |
| Comorbidity<br>(%) | 28,8 | 23,4 | 20 | 24,4 | 23,4 | 26,6 |
| Hipertension<br>(n) | 9 | 6 | 2 | 6 | 6 | 3 |
| Diabetes<br>Miellitus (n) | 3 | 3 | 0 | 4 | 3 | 1 |
| Respiratory<br>Disease (n) | 0 | 2 | 1 | 1 | 0 | 0 |
| Others (n) | 1 | 0 | 0 | 0 | 2 | 0 |

NEA- non-endemic area; EA- endemic area; NC- negative control; FS- flu syndrome; BMI – body mass index calculated by the formula $\text{kg/m}^2$ where kg is a person's weight in kilograms and $\text{m}^2$ is their height in squared meters.

Volunteer recruitment spanned from December 2020 to October 2021, focusing on identifying individuals displaying symptoms akin to flu-like syndromes of unknown origin (Flu-like syndrome) and COVID-19, alongside including individuals in good health (control group). Throughout the collection period, circulating strains of COVID-19 encompassed both the original variant and P1. All chosen volunteers underwent an RT-PCR (Real-Time Polymerase Chain Reaction) test to validate the diagnosis of COVID-19. Volunteers were recruited at UPA-Centro Sul and Hospital Universitário Risoleta Tolentino Neves in Belo Horizonte and Hospital Unimed in Governador Valadares.

The study’s inclusion criteria involved recruiting adult or elderly volunteers aged 20 years old or above, irrespective of whether they exhibited flu-like symptoms, provided they consented to sign the Informed Consent Form (TCLE). The participant selection process also adhered to specific exclusion criteria, excluding children and adolescents (under 18 years old), individuals with inconclusive results in the RT-PCR test for COVID-19 detection, flu-like symptoms persisting beyond 9 days, and individuals living with HIV.

The body mass index (BMI) of the participants was categorized based on the following criteria: less than 18.5 (underweight), between 18.5 and 24.9 (normal weight), between 25.0 and 29.9 (overweight), and exceeding 30.0 (obesity).

The volunteers were engaged by the study team in health units where they reviewed and addressed any queries regarding the Informed Consent Form (ICF), they underwent a clinical and sociodemographic questionnaire session and had their blood samples collected for serum and plasma separation, alongside swab collection for SARS-CoV-2 detection via RT-PCR. Individuals diagnosed with COVID-19 were telemonitored for 14 days following confirmation and delivery of the RT-PCR test results.

### Luminex-Multiplex measurement of inflammatory mediators

To assess blood mediators, we utilized heparinized plasma from our volunteers, employing the Bio-Plex® Pro Human Cytokine Standard multiplex kit by Bio-Rad Laboratories. This kit facilitates the simultaneous analysis of numerous analytes through magnetic immunoassay, conducted on Luminex equipment (Bio-Plex® 200, Bio-Rad). The quantified panel of analytes encompasses IL-1β, IL-1Ra, IL-2, IL-4, IL-5, IL-6, IL-7, CXCL8, IL-9, IL-10, IL-12p70, IL-13, IL-15, IL-17A, CCL11, FGF-Basic, G-CSF, GM-CSF, IFN-γ, CXCL10, CCL2, CCL3, CCL4, PDGF-BB, CCL5, TNF-α, and VEGF (Table 4). The analyses were conducted utilizing the Bioplex™ xPONENT software version 3.1 by Bio-Rad.

### Radar plots

The radar chart depicted cytokines/chemokines categorized as low (≤ global median) and high (≥ global median) producers, utilizing the global median of each mediator as a cut-off (Silveira-Nunes et al., 2017). The calculation of the global median encompassed the entire dataset derived from the respective groups. Within the radar, each axis symbolizes individuals exhibiting elevated levels of a specific mediator (termed high producers). These axes interconnect to form a polygonal area, illustrating the equilibrium between growth factors, inflammatory, and anti-inflammatory mediators. An increase or decrease in this area signifies a higher or lower contribution of each mediator to the overall profile.

### DNA extraction and bisulphite treatment

Genomic DNA extraction was conducted from peripheral whole blood samples using the Trizol and Chloroform protocol. Subsequently, 1 μg of DNA underwent bisulfite conversion utilizing the EZ-96 DNA Methylation Kit (Zymo Research, Irvine, USA) with specific adjustments: incubation in CT buffer for 21 cycles of 15 minutes at 55°C and 30 seconds at 95°C, followed by elution of the bisulfite-treated DNA in 100 μl of water. Post-extraction and conversion, the samples were loaded onto the Illumina Infinium Methylation EPIC Bead Chip. Fluorescence data were collected from all enrolled subjects, and subsequent beta values were calculated.

### Estimation of DNAm age

Following the acquisition of the methylation beta value matrices file, we utilized the methylclock 1.5.0 package (Pelegi-Siso et al., 2021) to estimate biological ages and aging acceleration through nine distinct methylation clocks.

### Serology

Serological analysis was carried out on individuals from EA (Governador Valadares, Brazil) to detect IgG antibodies against the dengue virus, cytomegalovirus (CMV), and the anti-K39 leishmaniasis antigen. All examinations were outsourced to the Hermes Pardini Laboratory in Belo Horizonte, Brazil, adhering to the institution’s protocols for detection techniques, sample transportation, and storage methods. Dengue serology employed Enzyme Immunoassay, facilitating the quantitative detection of specific IgG for all dengue serotypes (DEN1, 2, 3, and 4). CMV testing utilized the ELFA (Fluorimetry Enzyme Assay) technique, allowing the quantitative detection of IgG antibodies specific to CMV types 1 and 2 antigens. Lastly, leishmaniasis serology was performed using Immunochromatography, enabling the detection of the K-39 antigen, widely utilized in confirming suspected cases of Visceral Leishmaniasis and chronic instances of the disease.

### Spearman’s correlation analysis

Spearman’s coefficient analysis was performed using the R package to assess multiple associations between biomarkers and determine the strength of relationships. The correlation coefficient value ranges from −1 to +1, where a positive value indicates changes in the same direction and a negative value indicates opposite directions. A value of 0 signifies no association between variables. The correlation plot displays only those correlations with p < 0.05. Circle size represents statistical significance, with larger circles denoting lower p-values and increased significance.

### Statistical analysis

The significance of differences between groups was assessed through parametric tests (Student’s t-test and analysis of variance – ANOVA, followed by Tukey’s post-test) or non-parametric tests (Mann-Whitney U and Kruskal-Wallis test, followed by Dunn’s post-test). The normality of sample distribution was assessed using the Kolmogorov-Smirnov test. Participant characteristics were presented as mean (standard deviation) or median (minimum to maximum range) for continuous variables and as frequency (number) for categorical variables. When appropriate, means (standard deviation) or medians (interquartile range) were depicted in graphs. All statistical analyses were conducted using the GraphPad Prism 8.0 package, considering a difference as significant when p < 0.05.

## Results

### Individuals Residing in Endemic Areas had Accelerated Epigenetic Aging

A previous report conducted by our group described that individuals living in an area endemic for infectious diseases had accelerated biological age measured by DNA methylation(Durso et al., 2022). This study included volunteers from Governador Valadares, MG, an endemic area (EA), and individuals from São Paulo, SP, a non-endemic area (NEA). In this study, we also compared the epigenetic aging of the volunteers from another city, Belo Horizonte, MG, as a NEA sample. Epigenetic analyses indicated that individuals living in EA also exhibited accelerated aging compared to those living in NEAs across the six biological clocks evaluated: skinHovarth (Fig. 1a), PedBE (Fig. 1b), Wu (Fig. 1c), TL (Fig. 1d), BLUP (Fig. 1e), and EN (Fig. 1f).

**Fig 1:**
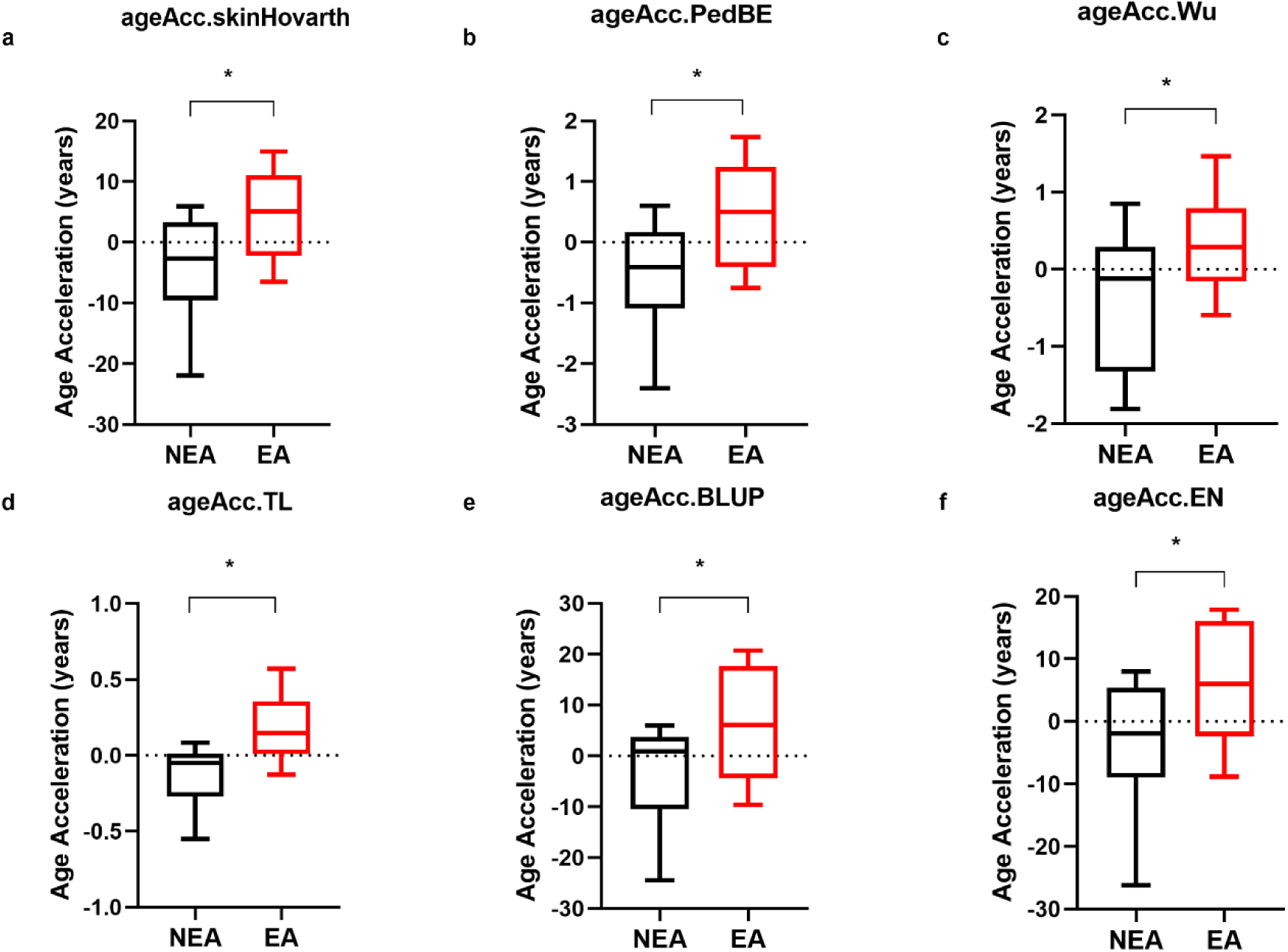

### Individuals Residing in Endemic Areas for Infectious Diseases Presented Higher Production of IL-12p70, IL-17A, and IL-9

Since aging can lead to various alterations in the immune response (Franceschi et al., 2007), we investigated the immune repercussions in the production of inflammatory mediators in our cohort. Residents in EA had higher production of IL-12p70 (Fig. 2a), IL-17A (Fig. 2b), and IL-9 (Fig. 2c) compared to individuals living in NEA. Of note, these individuals were reactive to other viral infections, such as dengue and CMV (see Table 2). On the other hand, residents in NEA showed higher production of IL-6 (Fig. 2d), IL-1β (Fig. 2e), IL-2 (Fig. 2f), IL-1ra (Fig. 2g), and IL-10 (Fig. 2h). Additionally, individuals residing in EA presented a less homogeneous profile and a higher frequency of high producers of PDGF-BB, VEGF, CXCL11, IL-9, IL-12p70, and IL-17A compared to NEA residents (Fig. 2i, j).

**Fig 2:**
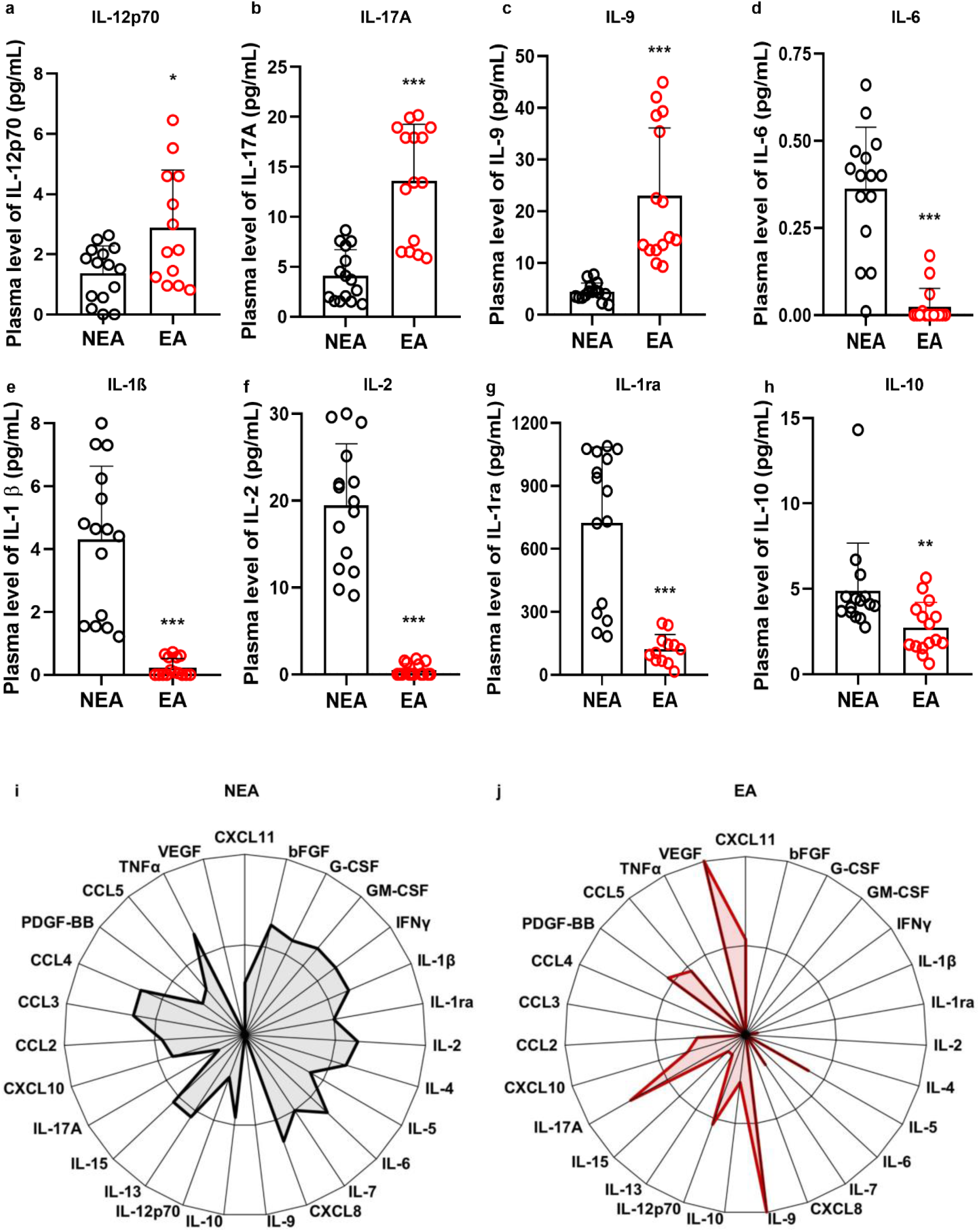

**Table 2.** Prevalence of CMV, Dengue, and Leishmania infections among negative control (NC) individuals residing in the EA based on serological analysis:

| **Negative control individuals residing in EA (n=15)** |  |
| --- | --- |
| CMV | 100% |
| Dengue | 100% |
| Leishmania | 0% |

### Individuals from endemic area with either Flu-Like Symptoms or COVID-19 had Higher Production of plasma mediators

Alterations in immune activity associated with aging can increase vulnerability to infectious diseases (Bernabeu-Wittel et al., 2020). In this study, we explored putative differences in the production of inflammatory mediators in response to viral infections between the two cohorts. Participants from endemic areas for infectious diseases (EA) were compared to those from non-endemic areas (NEA) who exhibited either flu-like symptoms or COVID-19. Individuals with either flu-like symptoms or infected with COVID-19 residing in EA exhibited higher production of IL-12p70 (Fig. 3a), IL-6 (Fig. 3b), IL-1β (Fig. 3c), IL-2 (Fig. 3d), and IL-1ra (Fig. 3e) compared to NEA residents. Additionally, EA individuals with COVID-19 showed increased production of IL-10 compared to NEA individuals (Fig. 3f).

**Fig 3:**
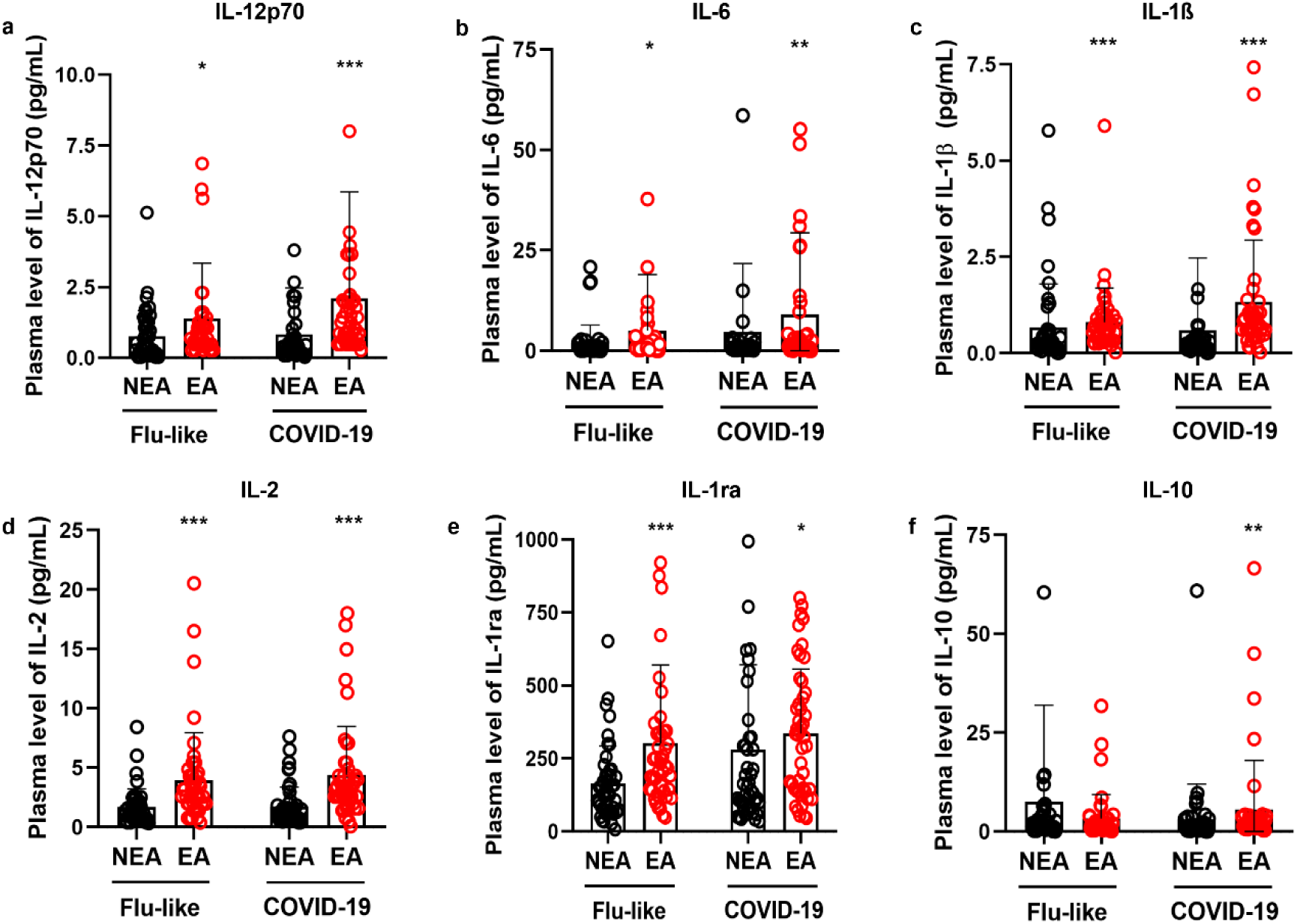

### High Producer Profile of Individuals Residing in Endemic Areas versus Non-Endemic Areas

In addition to the differences found in cytokine production, we observed that these individuals exhibited a higher frequency of high producers of inflammatory mediators, anti-inflammatory mediators, chemokines, and growth factors compared to NEA residents, both with flu-like syndromes (Fig. 4a, b) and COVID-19 (Fig. 4c, d). Furthermore, the inflammatory profile of individuals residing in EA is very similar to the profile of individuals residing in NEA regardless of the type of infection, (Fig. 4c, d).

**Fig 4:**
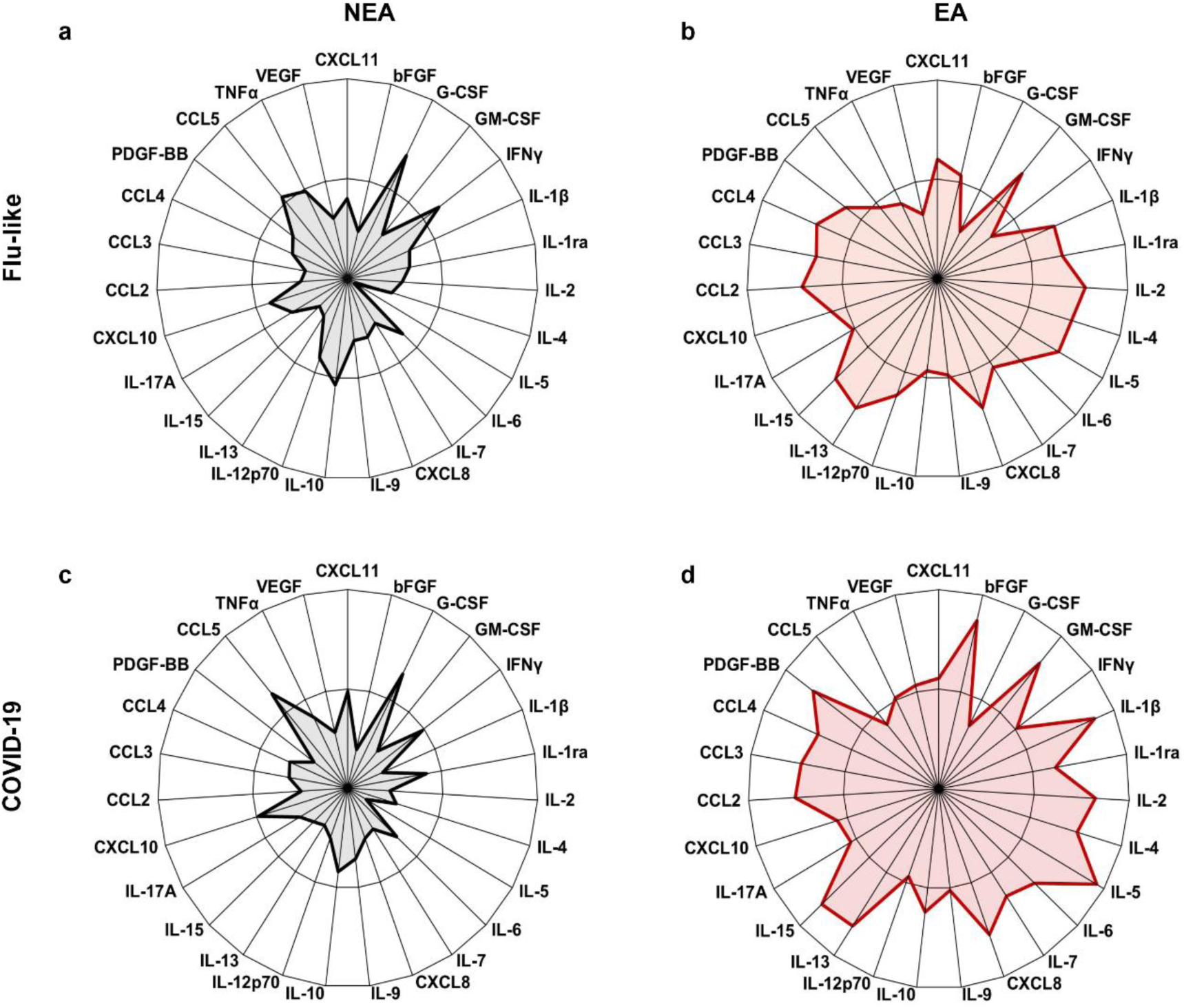

### Individuals Residing in Endemic Area had Distinct Correlations Among Inflammatory Mediators **a**nd Epigenetic Age

Aging is accompanied by a state of low-grade chronic inflammation, known as “inflammaging“(FRANCESCHI et al., 2000). Since individuals residing in EA exhibit accelerated aging and possess a distinct profile in the production of inflammatory mediators compared to NEA residents, we aimed to understand the biomarkers associated with the biological aging of these individuals by comparing correlation matrices. The correlogram of individuals living in EA showed more intense positive correlations among inflammatory mediators than the correlogram of individuals living in NEA (Fig. 5 a, b). Furthermore, epigenetic age is positively correlated, in increasing order, with CXCL8, CCL5, CCL4, IL-5, CCL3, TNFα, IL-17A, CXCL11, IL-9, IL-6, bFGF, CXCL10, G-CSF, IL-7, CCL2, IL-4, VEGF, and GM-CSF in EA volunteers, while in NEA volunteers, epigenetic age is positively correlated, in increasing order, with IL17A, CCL2, IL-4, IL-10, CXCL11, IL-12p70, IL-7, CCL5, and IL-9 (Fig. 5 d).

**Fig 5:**
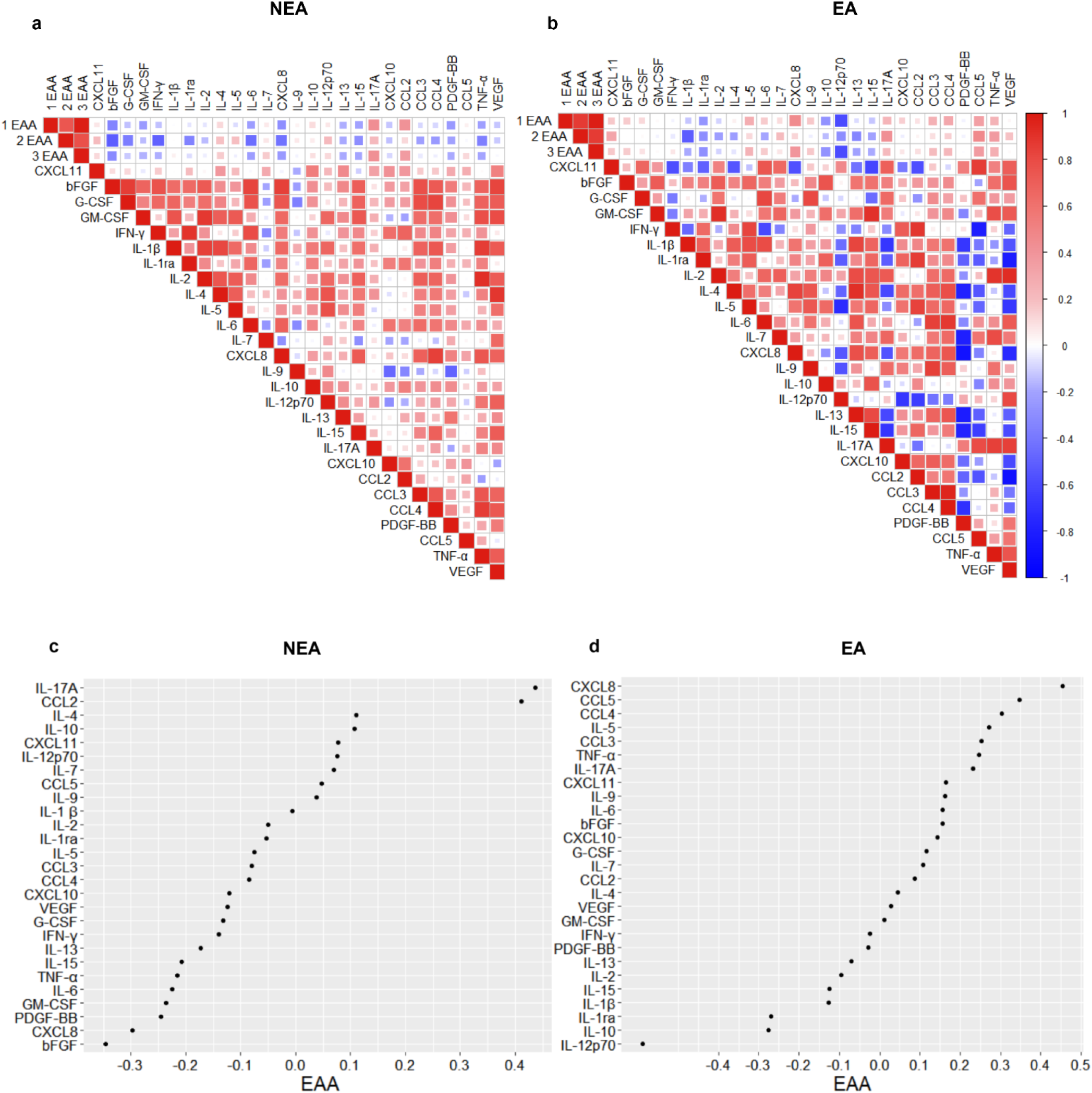

## Discussion

Environmental factors, such as lifestyle, can play a crucial role in promoting healthy aging. While chronological aging progresses at a fixed rate, the rate of biological aging varies among individuals. Biological age can be measured by epigenetic clocks, which are based on DNA methylation patterns (Field et al., 2018).

To understand whether frequent exposure to infectious antigens leads to accelerated aging, we used six epigenetic clocks to compare the aging of individuals residing in an EA with those in a NEA. Our results showed that individuals in the EA exhibit accelerated epigenetic aging across all evaluated methylation clocks (Fig. 1a-f). These findings are consistent with results from a previously published study by our group, despite using a cohort from a different NEA(Durso et al., 2022).

Among the various modifications associated with aging, numerous changes in immune activity are observed. One of the most prominent is the “inflammaging”, a chronic low-grade inflammation present in the elderly, characterized by the production of IL-1, IL-6, and TNF-α(FRANCESCHI et al., 2000). In this study, differently from the previous one, we examined the production of inflammatory mediators in our two cohorts of individuals and found that residents in EA had increased production of IL-12p70 (Fig. 2a), IL-17A (Fig. 2b), and IL-9 (Fig. 2c). In contrast, individuals residing in the NEA showed higher production of IL-6 (Fig. 2d), IL-1β (Fig. 2e), IL-1ra (Fig. 2f), IL-2 (Fig. 2g), and IL-10 (Fig. 2h).

Although our results indicate higher production of IL-6 (Fig. 2d) and IL-1β (Fig. 2e) in NEA residents—cytokines typically associated with *inflammaging*, a phenomenon commonly linked to elderly populations—it is important to note that the volunteers in both groups were age-matched. Furthermore, this study does not focus on a cohort of elderly individuals but rather on two cohorts with distinct biological aging profiles. It is worth emphasizing that aging is a continuous process that occurs throughout life, beginning at birth.

IL-12 consists of the p35 and p40 subunits, which combine to form the bioactive IL-12p70. This cytokine is crucial for IFN-γ production and the activation of Th1 cells(Gee et al., 2009). IL-17A, on the other hand, is primarily produced by Th17 cells in response to IL-1β and IL-23, mediating immunity against fungi and bacteria(Mills, 2023). IL-9 is a pleiotropic cytokine produced by Th2 cells, ILC2s, mast cells, and basophils, playing a significant role in type 2 immunity, autoimmunity, and the immune response to tumors(Pajulas et al., 2023). IL-6 is an acute-phase pro-inflammatory cytokine secreted primarily by macrophages, important for regulating inflammatory disorders such as viral infections(Aliyu et al., 2022; Gubernatorova et al., 2020). IL-1β, also produced by macrophages, is cleaved by inflammasome activation, which is triggered by caspase-1. Besides protecting against pathogens, this cytokine regulates sterile insults and plays a crucial role in proliferation, differentiation, and apoptosis (Bent et al., 2018). Conversely, IL-1ra acts as a natural inhibitor of IL-β by serving as an antagonist of the interleukin-1 receptor(Yazdi and Ghoreschi, 2016). Increased production of IL-1ra is a regulatory response to control the elevated secretion of IL-1β. IL-10 exerting potent immunosuppressive functions(Wang et al., 2019). On the other hand, IL-2 can have opposing functions playing a significant role in both inflammation and suppression. It is involved in the proliferation of T cells, facilitating the generation of effector and memory cells, and in the maintenance of regulatory T cells(Abbas et al., 2018).

In response to immunogenic challenges, the immune response typically polarizes towards Th1, Th2, or Th17 pathways. Interestingly, our data revealed a lack of polarization in the profile of mediators from individuals residing in EA with inflammatory cytokines exerting diverse functions. In contrast, the panel of mediators produced by individuals in NEA was more balanced, with elevated production of both early inflammatory cytokines and anti-inflammatory cytokines. This suggests that at steady state individuals residing in endemic areas had an immunological plasma profile very distinct from the one exhibited by individuals from non-endemic areas.

Inflammation is a natural process that plays a crucial role in various functions, including embryogenesis and tissue repair (Ramos, 2012). A balanced profile of immune response is considered a marker of health since chronic inflammation is detrimental and can contribute to biological aging, as observed in studies of individuals with HIV (Giron et al., 2024), autoimmune diseases such as systemic lupus erythematosus(Mazzone et al., 2019), and conditions like obesity or type 2 diabetes(Ling and Rönn, 2019). Despite the differences observed in the production of inflammatory mediators between the groups, the levels remained subclinical, and no symptom was reported. Moreover, participants in both groups do not exhibit significant comorbidities or differences in age or sex (Table 1). However, all individuals residing in EA tested positive for viral infections, such as dengue and CMV, in serological tests (Table 2). It is likely that these infections impacted in the inflammatory profile of individuals form EA(Pawelec, 2014).

It is known that changes in immune activity associated with aging can increase vulnerability to diseases (Bernabeu-Wittel et al., 2020; Chung et al., 2018). Therefore, we also investigated whether individuals residing in endemic areas exhibited differences in the production of inflammatory mediators in response to viral infections. We compared residents of EA with those in NEA who had flu-like symptoms or were diagnosed with mild COVID-19 according to the World Health Organization (WHO, 2020) classification criteria.

Residents of EA with COVID-19 and FLS had higher production of IL-12p70, IL-6, IL-1β, IL-2, and IL-1ra compared to residents of NEA (Fig. 3a-e), as well as an increased production of IL-10 in COVID-19 patients (Fig. 3f). Additionally, these individuals showed a greater frequency of high producers of cytokines, chemokines, and growth factors, compared to those in NEA, both among those with flu-like symptoms (Fig. 2a, b) and those with COVID-19 (Fig. 2c, d).

Therefore, individuals residing in EA not only responded more intensely to infections than those in NEA, but they also produced a distinctive panel of mediators during these responses. We hypothesize that this may be due to a combination of an immunosenescent inflammatory profile, characteristic of those exposed to multiple infections throughout life (Fig. 2), and their response to the viral antigens they encountered at the time of the study. A previous report supports this hypothesis, as it demonstrated that even young adults co-infected with cytomegalovirus and Epstein-Barr virus already had an aging-related T cell phenotype (Hofstee et al., 2023).

Additionally, our results showed that although individuals with COVID-19 residing in EA produced higher levels of mediators compared to those with flu-like symptoms (Fig. 4c, d), the inflammatory profile of EA residents was very similar regardless of the type of infection (Fig. 4b, d). The same was true for residents in NEA (Fig. 4a, c).

These results lead us to believe that environmental differences, triggered by local stressors, were associated not only with varying rates of biological aging but also with distinct immunological profiles at steady state and during viral infectious diseases.

To understand whether there are differences in aging-associated inflammatory mediators between individuals from EA and those from NEA, we correlated three epigenetic clocks with the production of inflammatory mediators. The correlation matrix for individuals living in EA showed a greater number of positive correlations and stronger correlations among inflammatory mediators compared to the correlogram for individuals in NEA (Fig. 5a, b).

Additionally, epigenetic age was positively correlated, in descending order, with CXCL8, CCL5, CCL4, IL-5, CCL3, TNF-α, IL-17A, CXCL11, IL-9, IL-6, bFGF, CXCL10, G-CSF, IL-7, CCL2, IL-4, VEGF, and GM-CSF among EA volunteers (Fig. 5d). In contrast, for NEA volunteers, epigenetic age was positively correlated, in descending order, with IL-17A, CCL2, IL-4, IL-10, CXCL11, IL-12p70, IL-7, CCL5, and IL-9 (Fig. 5c).

It has been proposed that aging is not characterized solely by deleterious changes and functional decline, but it includes a remodeling process to cope with challenges that occurs throughout life(Raffington, 2024). In this sense, aging involves compensatory changes resulting from the immunological history of individuals. Our data on the immunological and biological age of individuals who are residents of EA and NEA confirm this hypothesis.

Several studies indicate that aging can be accelerated by various lifestyle factors, such as processed foods and poor sleep quality(Friedman, 2020). In this study, we added a layer of complexity to these findings demonstrating that living in endemic areas for infectious disease may also impact on the immunological profile of individuals and in their aging process. Continuous exposure to strong antigenic stimulation is probably determinant for these differences, and they may be an important factor for the aging process of populations from many regions of the world, not only Brazil. Therefore, these data are relevant for a more universal understanding of immunosenescence and for the design of strategies to promote healthy aging.

## Author contributions

AMC designed the study, coordinate the recruitment and all the analysis of the project, wrote the manuscript; M.M.C. designed the work, performed most of analysis and wrote the manuscript; F.C.M. performed the bioinformatic analysis and discussed the results; L.W.Z. helped with the DNA methylation analysis and the discussion of the results; L.T. and L.H.A.V. performed the Luminex analysis and helped with discussion of the results; L.H.A.V. and G.C.C. performed the recruitment and blood collection of the volunteers; B.S-S. helped with the DNA methylation analysis and the discussion of the results; D.F.D. helped with the analysis of DNA methylation data; H.I.S. performed the RT-PCR tests of all volunteers under the supervision of S.M.R.T.; M.S.C. performed the clinical exams of volunteers at UPA-Centro Sul in Belo Horizonte under the supervision of U.T.; H.C.G. and R.C.B. were in charge of clinical exams of volunteers recruited at Hospital Universitário Risoleta Tolentino Neves in Belo Horizonte; M.L.O.J. was in the charge of the clinical exams of volunteers recruited at Hospital Unimed in Governador Valadares; E.S.F. helped with the Luminex analysis and analysis of the data; G.S-N. coordinate all the recruitment and sample collection of volunteers from Governador Valadares and helped with the statistical analysis; A.T-C. supervised the Luminex analysis, serology tests and helped with the discussion of the data; T.U.M. coordinate the recruitment, questionnaire application and telemonitoring of volunteers of Belo Horizonte.

## Ethical approval

The study complied with ethical guidelines established by CONEP, ensuring participant safety and well-being. It received approval from the Human Research Ethical Board of UFMG and CONEP (CAAE #40208320.3.1001.5149). Participants were fully informed about the study’s objectives and procedures and provided written consent before participating.

## Additional information

Correspondence and requests for materials should be addressed to A.M.C.

## Data availability

No datasets were generated or analysed during the current study.

## Acknowledgements

Not applicable.

